# Genomic epidemiology reveals multiple introductions and spread of SARS-CoV-2 in the Indian state of Karnataka

**DOI:** 10.1101/2020.07.10.20150045

**Authors:** Chitra Pattabiraman, Farhat Habib, Pk Harsha, Risha Rasheed, Vijayalakshmi Reddy, Prameela Dinesh, Tina Damodar, Kiran Hosallimath, Anson K George, Nakka Vijay Kiran Reddy, Banerjee John, Amrita Pattanaik, Narendra Kumar, Reeta S Mani, Manjunatha M Venkataswamy, Shafeeq K Shahul Hameed, B.G Prakash Kumar, Anita Desai, Ravi Vasanthapuram

**Author notes:** Corresponding author. Email –.

## Abstract

Karnataka, a state in south India, reported its first case of Severe Acute Respiratory Syndrome Coronavirus 2 (SARS-CoV-2) infection on March 8, 2020, more than a month after the first case was reported in India. We used a combination of contact tracing and genomic epidemiology to trace the spread of SARS-CoV-2 in the state up until May 21, 2020 (1578 cases). We obtained 47 full genomes of SARS-CoV-2 which clustered into six lineages (Pangolin lineages—A, B, B.1, B.1.1, B.4, and B.6). The lineages in Karnataka were known to be circulating in China, Southeast Asia, Iran, Europe and other parts of India and are likely to have been imported into the state both by international and domestic travel. Our sequences grouped into 12 contact clusters and 11 cases with no known contacts. We found nine of the 12 contact clusters had a single lineage of the virus, consistent with multiple introductions and most (8/12) were contained within a single district, reflecting local spread. In most of the twelve clusters, the index case (9/12) and spreaders (8/12) were symptomatic. Of the 47 sequences, 31 belonged to the B/B.6 lineage, including seven of eleven cases with no known contact, indicating ongoing transmission of this lineage in the state. Genomic epidemiology of SARS-CoV-2 in Karnataka suggests multiple introductions of the virus followed by local transmission in parallel with ongoing viral evolution. This is the first study from India combining genomic data with epidemiological information emphasizing the need for an integrated approach to outbreak response.

## Introduction

Severe Acute Respiratory Syndrome Coronavirus 2 (SARS-CoV-2), a novel coronavirus that was first detected in individuals with acute pneumonia in China in late 2019, has now spread throughout the world^1^. The World Health Organization (WHO) on March 11 declared the disease Coronavirus Disease 2019 (COVID19) caused by SARS-CoV-2 a pandemic^2^. COVID19 has claimed over 500,000 lives (as of July 5, 2020) and the pandemic is ongoing^3^.

Genomic epidemiology from the analysis of viral sequences from all over the world is consistent with the emergence of the virus in late 2019 in China and consequent spread and expansion in Europe, and other parts of the world^4,5^. More than 50,000 complete genomes of SARS-COV-2 from all over the world are currently available in public databases such as the GISAID initiative (originally known as Global Initiative on Sharing All Influenza Data)^6^. While this information is invaluable for understanding evolution of the virus^7^, pathogenesis, and design of diagnostic tools, few studies have been able to combine this with epidemiological data to derive insights on viral spread. These studies have provided information on how the virus is introduced and spread in a population.

A comprehensive study of circulating variants of the virus in Iceland, which included over 580 complete genomes in combination with epidemiological information (travel history and contact tracing) revealed that while the initial importation of the virus was from China and Southeast Asia subsequent importations were from different parts of Europe^8^. Studies based on complete SARS-CoV-2 genomes from Guangdong Province in China highlighted the initial importation of virus to the province by travel and limited local transmission^9^. A study of viral genomes from the east coast of the USA combined with travel data revealed the coast-to-coast spread of virus within the country^10^. Initial studies in Washington State uncovered cryptic local transmission^11^. These studies reiterate the importance of combining sequencing data with public health information.

The first case of COVID19 in India was detected on January 30, 2020 and case numbers have continued to rise inspite of stringent interventions including nationwide lockdowns. In the first few months of the outbreak, between January 22–April 30, 2020, test results from all over India could be averaged to a positivity rate of 3.9%^12^. Analysis of these cases revealed that the test positivity rate was highest when the samples were from contacts of a known COVID19 positive case^12^.

A large number of SARS-CoV-2 genomes (about 1500 complete genomes as of July 5, source— GISAID) have been sequenced in different parts of India. The first sequences from India were reported from individuals with travel history to China, Italy and Iran^13,14^.

An analysis of 361 complete genome sequences from India showed that five global clades were circulating in India – old Nextstrain clades B, B4, A2a, A3, and a distinct clade A3i^15^. The A2a (European) clade was found to be the most prevalent, followed by A3i^15,16^. While these studies have added valuable information on circulating lineages of SARS-CoV-2 in India, they have not comprehensively linked genomic data with epidemiological information. This study was therefore undertaken to dissect the molecular epidemiology of SARS-CoV-2 in Karnataka, a state in South India. Here we report 47 full-length SARS-CoV-2 genome sequences obtained from individuals who tested positive for the virus by RT-PCR and present an analysis of epidemiological information combined with genomic data to elucidate the introduction and spread of the virus in the state.

## Methods

### Samples and epidemiological data

Samples received at the National Institute of Mental Health and Neuroscience (NIMHANS), Bengaluru for COVID19 RT-PCR testing between March 5–May 21 2020, were included in this study. This study was approved by Institutional Ethics Committee (Basic and Neurosciences). The line list of positive patients was provided by the Directorate of Health and Family Welfare Services, Karnataka and missing data was filled in from the ICMR (Indian Council of Medical Research) portal. The line list contained detailed epidemiological workup of each sample including information on age, sex, clinical signs and symptoms, location, contacts, description of exposure type, sampling dates, date of hospitalization and follow up of hospitalized cases.

### Amplicon Sequencing and recovery of SARS-CoV-2 genomes

Samples received for testing at NIMHANS were subjected to RT-PCR based on ICMR guidelines^17^, details of the kits used are provided in Supplementary Table 1. A total of 21,418 samples were tested in NIMHANS (April 5, 2020–May 20, 2020), 369 of these were positive and 60 were included for sequencing. The criteria for sequencing was RT-PCR positive samples with Ct values under 30. Samples with Ct value > 30 were included when they were considered critical for representing a cluster or if they were from symptomatic individuals.

We used a tiling primer based approach for whole genome sequencing described by the ARTIC Network using Primal Scheme^9,18^. Briefly, we used the V3 primer set—these are 96 pairs with amplicons of about 400 basepairs (bp) spanning the whole genome except 31bp of the 5’ and a part of the 3’UTR. PCR was performed by pooling adjacent/overlapping primers into different pools so as to prevent preferential amplification of short fragments between adjacent primer pairs. Primers were initially used at a concentration of 10μM as per the protocol, then modified to amplify regions that were missed by increasing primer concentrations to 50μM. For four regions additional primers were designed and a separate reaction was set up before the pooling step in order to complete the genome. The resulting PCR amplicons were used for preparing libraries for Nanopore sequencing using the native barcoding (NBD 104/114) approach combined with the ligation sequencing kit (SQK-LSK109). Between 12–24 samples were barcoded and included in a single run. The resulting DNA was cleaned up and added to FLO-MIN-106 flow cell and sequenced on the MinION.

### Analysis of sequencing data

Sequences were basecalled and demultiplexed using guppy (v3.6), read lengths between 100– 600bp were considered for further analysis. Primers were removed from the sequencing reads by trimming 25bp at the ends and additional trimming based on alignment using BBDuk (v1).

Resulting reads were mapped to the RefSeq strain (NC_045512) using Minimap2 (v1.17) within Geneious Prime (Geneious Prime 2020.0.3). A consensus was created for regions with coverage >10x using the 0% majority rule and corrected. Consensus was aligned to the reference to ensure the correct reading frame and annotation was transferred from the reference. Sequences were deposited into the GISAID database.

### Phylogenetic analysis, lineage assignment, detection of SNPs and amino acid replacements

Consensus sequences from the 47 genomes from this study were aligned with the reference genome using MUSCLE (v 3.8.425)^19^. The multiple sequence alignment was used to infer the phylogeny using iqtree (v 1.6.12)^20^. A total of 88 DNA models were tested, and the TPM2u+F+I model was selected based on the Bayesian Information Criterion. Maximum likelihood tree was constructed as the consensus tree from 1000 bootstraps, using the reference sequence (NC_045512) as the outgroup. Nodes with bootstrap values > 83% were used for interpretation. Time scaled phylogenies were constructed using TreeTime with the multiple sequence alignment described above and the date of sampling as dates. Pangolin lineage assignments were performed using the online tool^21^. Single nucleotide polymorphisms (SNPs) and amino acid replacements were detected using the CoV-Glue web application^22^. Both tools use sequences submitted to the GISAID database^6^.

### Analysis of epidemiological data and contact map

The epidemiological data was extracted from the line list and a contact map was constructed using the state line list of positive cases. We identified primary and secondary contacts for a patient from the line list. We then built a graph where each node is a positive individual and is connected by an edge with their contacts who were positive. This gives us the contact map. The graphs were then filtered by size of clusters or clusters containing a node with a particular property to focus on clusters of interest. The graphs are visualized using the d3.js^23^ library with attributes like clinical state, lineages, and geographical location indicated by colours of the nodes.

## Results

Karnataka recorded 1578 cases between March 5–May 21, 2020. Most of these cases were from six high burden districts, with Bangalore Urban (the district encompassing the city of Bengaluru) reporting 256 cases (Figure 1). In total 369 (23.38%) positives were recorded at our centre, of which 60 samples were taken for sequencing (Figure 1, Table 1). The features of positive cases in the Karnataka, and those tested and sequenced at our centre are in Table 1. Most of the positive individuals (1133/1578: 71.8%) in the state were below 40 years of age. More males than females tested positive (987/1578: 62.5%). Amongst the positive individuals in the state, 84.35% were asymptomatic (at sample collection). A total of 87.5% cases (1380/1578) had contact with a known COVID19 case or travel history. Amongst 369 positive cases tested at our centre, 168 did not have a known contact as of May 21, 2020. We included 11 of these 168 for sequencing.

**Table 1:** Characteristics of positive cases

|  | State | Our Centre |  |
| --- | --- | --- | --- |
|  |  | Lab | Sequenced |
| Number of positive cases** | 1578 | 369 | 60 |
| **Age** |  |  |  |
| 0-20 | 374 | 86 | 12 |
| 20-40 | 759 | 197 | 29 |
| 40-60 | 315 | 69 | 13 |
| 60-80 | 126 | 15 | 4 |
| 80-100 | 4 | 2 | 2 |
| **Sex** |  |  |  |
| Female (F) | 591 | 120 | 21 |
| Male (M) | 987 | 249 | 39 |
| M:F | 1.67 | 2.08 | 1.86 |
| **Clinical State** |  |  |  |
| Asymptomatic* (Asym) | 1331 | 351 | 52 |
| Symptomatic (Symp) | 247 | 18 | 8 |
| Ratio Asym*: Symp | 5.39 | 19.50 | 6.50 |
| **Nature of contact/ exposure** |  |  |  |
| Contact with COVID19 case | 700 | 201 | 43 |
| No known contact |  |  |  |
| i. Travel history (international) | 96 | 0 | 0 |
| ii. Travel history (domestic) | 584 | 104 | 6 |
| iii. ILI (under investigation) | 27 | 4 | 2 |
| iv. SARI (under investigation) | 55 | 5 | 3 |
| v. Under investigation | 62 | 27 | 3 |
| vi. Contact unknown | 54 | 28 | 3 |

\*\* Between 5 March – 21 May, 2020 \* Asymptomatic at sample collection
*Note: Nature of contact/exposure is classified as Contact with COVID19 case- where tested individual was in contact with a known positive case or No known contact, divided into six categories. i. Travel history (international) - travel history to other countries, ii. Travel history (domestic) - travel within the state or interstate, iii. ILI (under investigation) - individuals with Influenza like illness with no known source of infection, iv. SARI (under investigation) - individuals with severe acute respiratory infection where the source of infection is not known, v. Under investigation- source of infection is not yet known/ contact tracing has not been completed, vi. Contact Unknown- where the tested individual was from a location where there were cases (eg. A containment Zone) but a specific contact could not be identified.*

**Figure 1:**
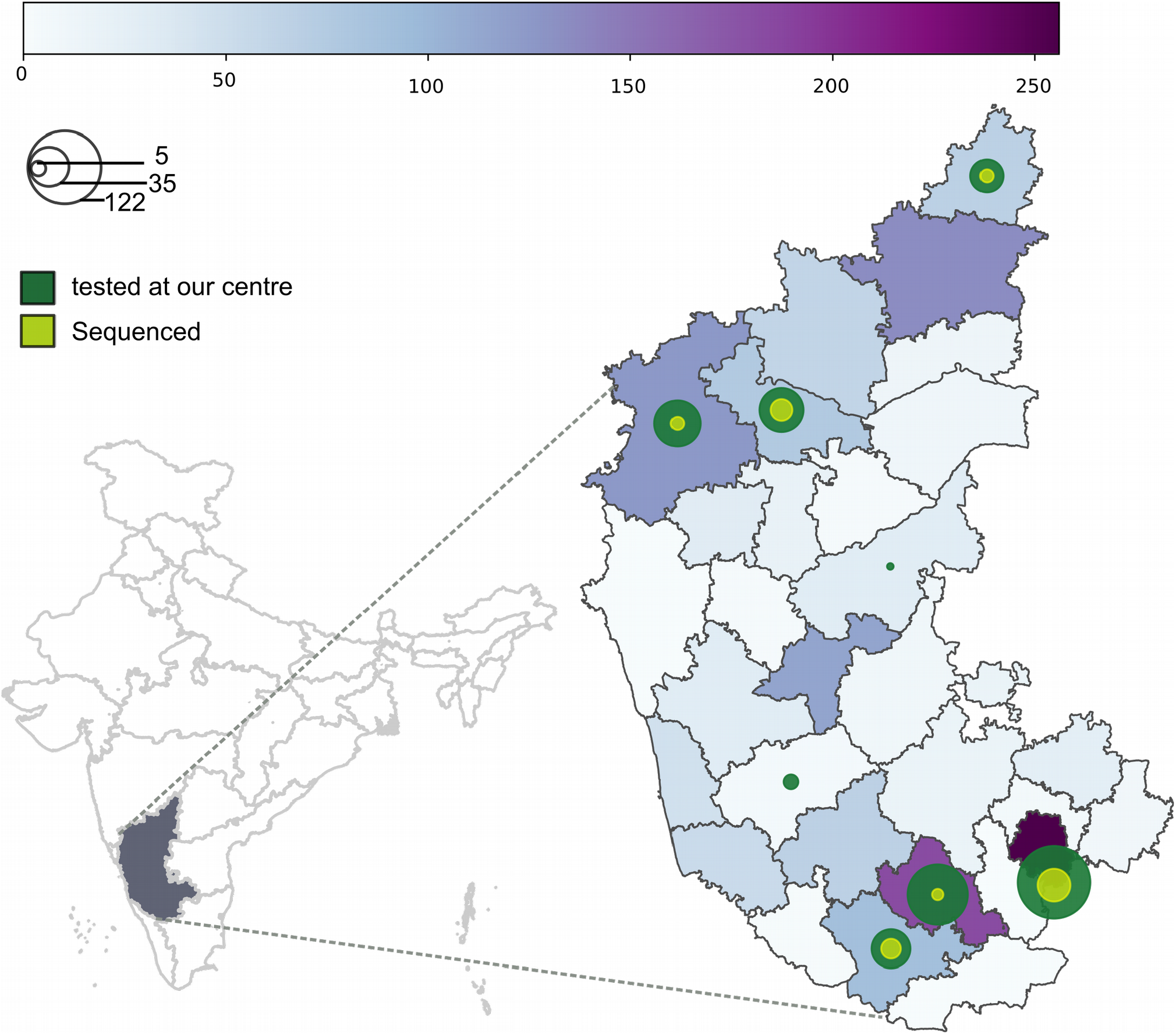
District wise distribution of SARS-CoV-2 positive cases in Karnataka sampled between March 5, 2020 to May 21, 2020. Left—Map of India highlighting the state of Karnataka. Right— Heat map shows the distribution of cases across 30 districts in Karnataka with high burden districts in deep purple. Size of the circle is proportional to the number of cases tested at our centre (green) and the number of cases sequenced (lime). The density of cases are represented by the heat map (horizontal bar) and concentric circles.

Overall 60 samples were sequenced, the Ct values of these samples ranged from 17.1–37.85, (Supplementary Table 1) and 47 complete genomes (1x coverage of 92.9–99.9%) were obtained. The average depth of coverage was over 3000x (Supplementary Figure 1). For the samples that yielded complete genomes about 200,000 sequences were obtained per sample, 89.3% (average) of the sequenced reads mapped to the reference genome (Supplementary Table 1, Supplementary Figure 1).

Lineage assignments using the Pangolin online tool revealed that the 47 complete genome belonged to six distinct lineages, A.p7 (3), B(14), B.1.1(3), B.1(8), B.6(17) and B.4(2) (Figure 2). Five of these lineages were apparent by maximum likelihood based phylogeny with bootstrap supports of >83%. Clades B and B.6 clustered together in this analysis. A time scaled maximum likelihood phylogeny of the genomes with the reference sequence shows that the lineages branched out at different time points (Figure 2) with defining mutations (Supplementary Figure 2) and that clade B.6 shared an ancestor with clade B more recently in time (Figure 2).

**Figure 2:**
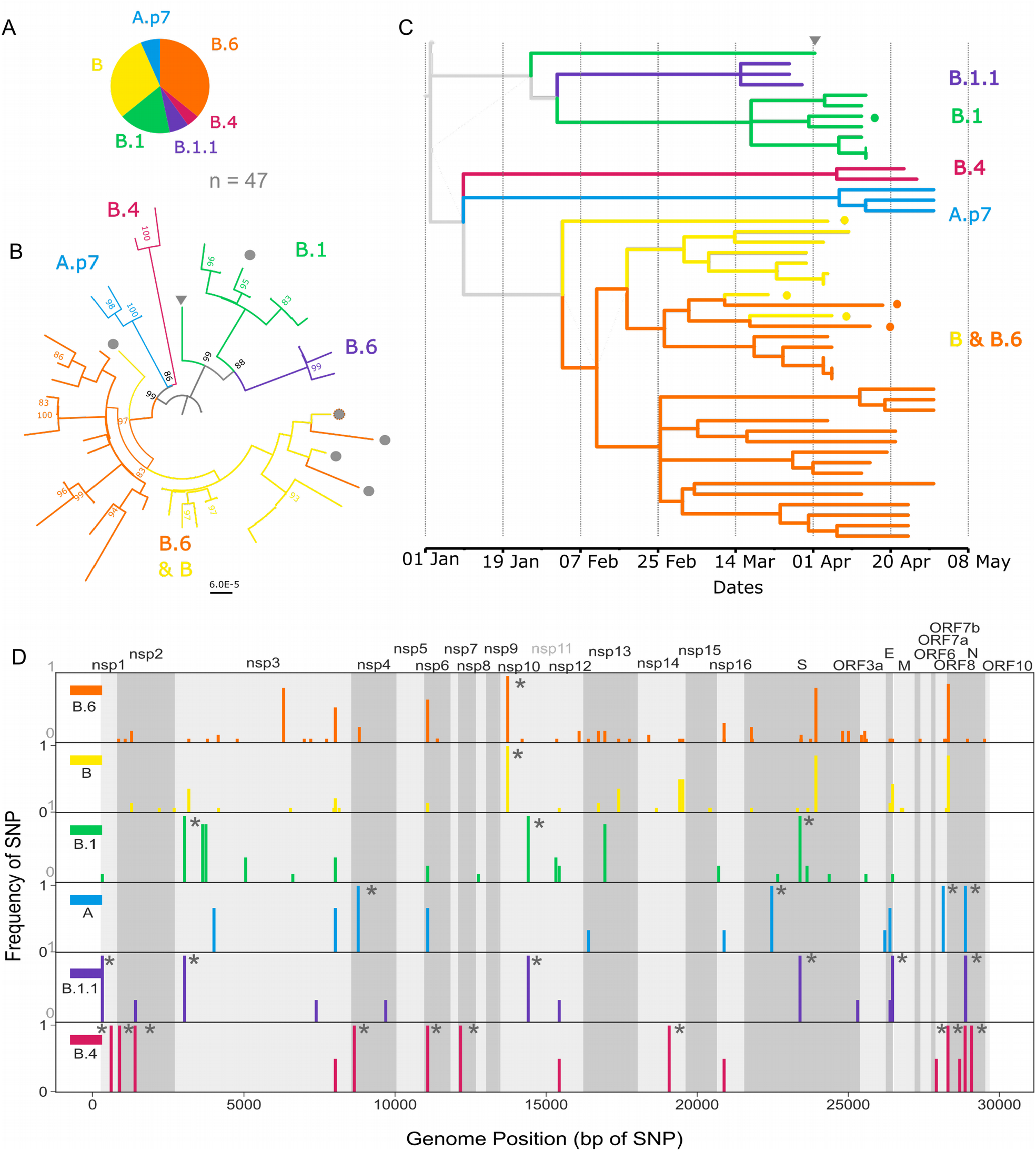
Six lineages of SARS-CoV-2 circulating in Karnataka. A. Pie chart shows the proportion of different lineages assigned to the 47 SARS-CoV-2 genomes from this study using the Pangolin tool. B. Maximum likelihood tree was constructed from 47 complete genomes using the reference SARS-CoV-2 genome (NC_045512) as the outgroup. Bootstrap support values over 83 are shown. Lineages are coloured based on Pangolin assignment. Grey circles indicate sequences from symptomatic individuals. C. Time scaled maximum likelihood tree of genomes from this study providing a chronology to introduction/importation events and propagation of the lineages post introduction into the state. Coloured circles represent sequences from symptomatic individuals. Grey arrowhead in B-C marks an outlier in the phylogeny from a case with no known contacts D. Position of single nucleotide polymorphisms across all six lineages is shown as SNP frequency (number of sequences from the lineage that have the SNP/total number of sequences in the lineage). Lineage defining SNPs are marked with *. Details of SNPs are provided in Supplementary Table 2.

Overall 112 Single Nucleotide Polymorphisms (SNPs) were identified in the 47 genomes in comparison to the reference sequence (Supplementary Table 2). Proportion and position of the SNPs are shown (Figure 2). A total of 79 amino acid replacements were identified (Supplementary Figure 2, Supplementary Table 3).

Amongst the 11 sequences from individuals with no known contact (Table 2, Figure 3), three clustered with lineage A.p7. These individuals had known travel histories to other parts of India. Of the remaining eight, two individuals with severe acute respiratory illness (SARI) and one with no known contacts clustered into Lineage B. One each of individuals with influenza like illness (ILI), known domestic travel history and unknown contact clustered with lineage B.6, while yet another individual with unknown contact clustered with lineage B.1 (Table 2, Figure 3).

**Table 2:** Lineage Assignments of positive cases with no known contacts

| Table 2: Lineage Assignments of positive cases with no known contacts |  |  |  |  |
| --- | --- | --- | --- | --- |
| Sr. No | Nature of contact/exposure | Lineage | Date of sample | District |
| 1 | Contact unknown | B.1 | 2020-04-12 | Mysuru |
| 2 | Contact unknown | B | 2020-04-14 | Belagavi/Others |
| 3 | SARI (under investigation) | B | 2020-04-15 | Bengaluru urban |
| 4 | SARI (under investigation) | B | 2020-04-16 | Bengaluru urban |
| 5 | ILI (under investigation) | B.6 | 2020-04-25 | Bengaluru urban |
| 6 | Domestic Travel | A | 2020-05-10 | Bagalkot |
| 7 | Domestic Travel | A | 2020-05-10 | Bagalkot |
| 8 | Domestic Travel | A | 2020-05-10 | Bagalkot |
| 9 | Domestic Travel | B.6 | 2020-05-10 | Bagalkot |
| 10 | Under Investigation | B.6 | 2020-05-10 | Bidar |
| 11 | Under Investigation | B.6 | 2020-05-10 | Bidar |

**Figure 3:**
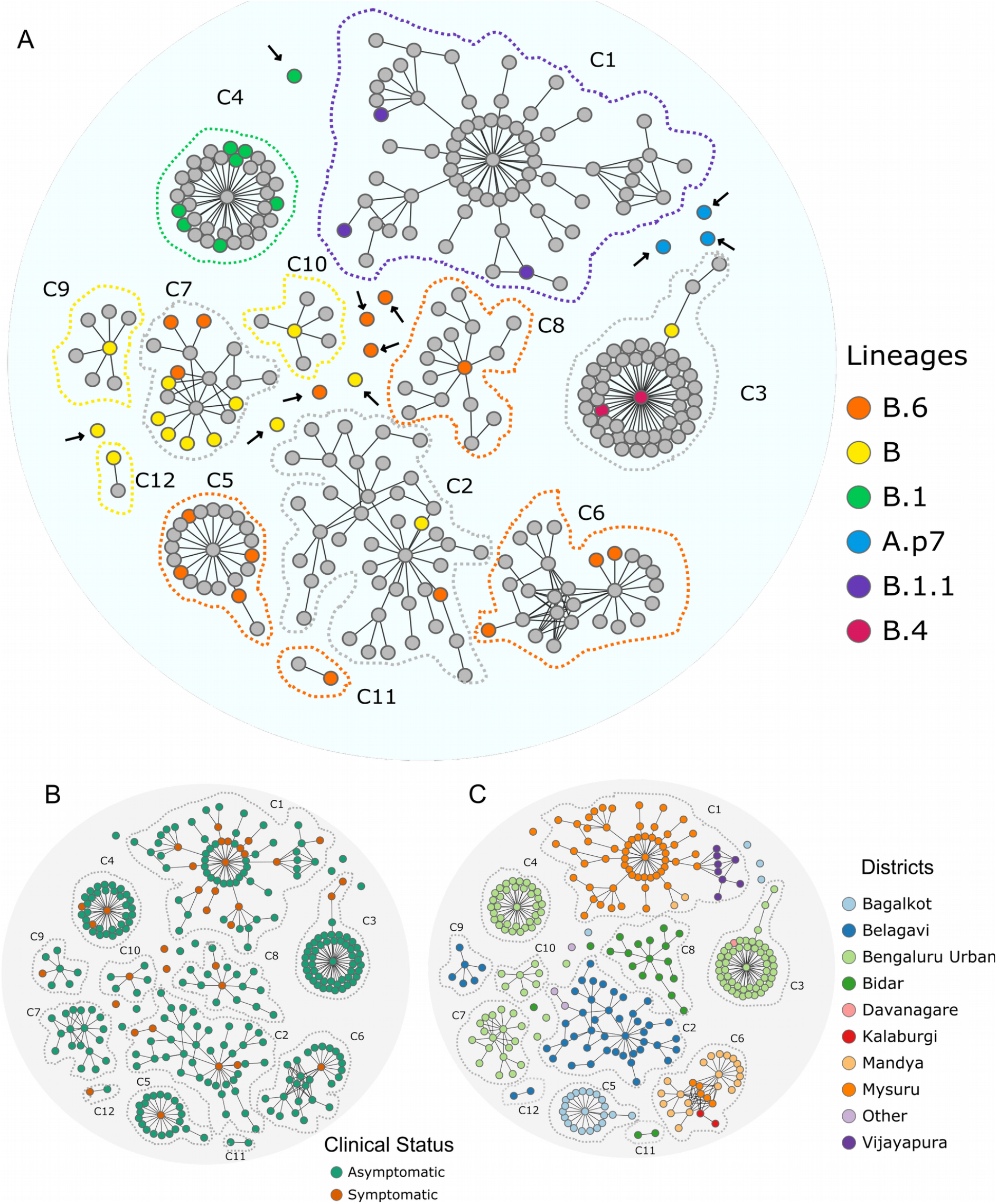
Contact graphs showing lineages, clinical state and geographical location of clusters. The graphs were made from analysis of contacts from the state line-list of cases and 47 sequences clustered into 12/104 clusters and 11 singletons (individuals with no known contact). These 12 clusters (C1-C12) and 11 singletons (n = 305 individuals) are shown in all the panels (A-C) **A**. Contact graph with individuals from whom complete genomes were obtained are coloured by lineage. Note: lineages were assigned to all 11 singletons (indicated with a black arrow) **B**. Contact graph of sequenced clusters and singletons are coloured by clinical status—symptomatic or asymptomatic. Red depicts symptomatic individuals and green represents the asymptomatic individuals. **C**. Graph representing geographic distribution of contact cluster by place of residence. Note: Most of the clusters are restricted to a district. A minority of cases (purple) are from districts other than those listed.

Analysis of contact information in the state line list, revealed that 822 of 1578 cases could be assigned into 104 contact clusters (Supplementary Table 4). Of these 104 clusters, 38 clusters were tested at our centre and 12 of the 38 were included for sequencing. These 12 included 294 people and covered ten large clusters (>5 individuals) from the state (Supplementary Table 4).

Of the 12 clusters (C1–C12), C1; C9–10, C12; C5–6, C8,C11; had sequences assigned to lineage B.1.1, B, B.6, and B.1 respectively. Three clusters had sequences assigned to more than one lineage. C2 and C7 had both lineage B and lineage B.6, C3 had sequences from lineage B.4 and lineage B (Figure 3).

Most of the cases in the state were asymptomatic (1331/1578) (Table 1, Figure 3). Analysis of information pertaining to the index cases (earliest detected individuals) from the 12 clusters revealed the following—nine of the 12 (75%) were symptomatic, of these six presented with SARI and the remaining three symptomatic had history of interstate travel (Supplementary table 4, Figure 3). Further, analysis of individuals with maximum number of contacts (spreader) within a cluster revealed that 8/12 were symptomatic (Supplementary Table 4).

The location of the clusters was analysed using a contact graph. Clusters (9/12) were limited to a single district excepting clusters C1, C3, C6, and C10 which were spread across districts. Time course of the 12 sequenced clusters and 11 cases with no known contacts indicated that i) lineage A.p7 was introduced recently ii) no new cases from C1 and C4 were detected (as of May 21, 2020) iii) ongoing transmission was evident for lineage B and lineage B.6. (Figure 4, Supplementary Table 5).

**Figure 4:**
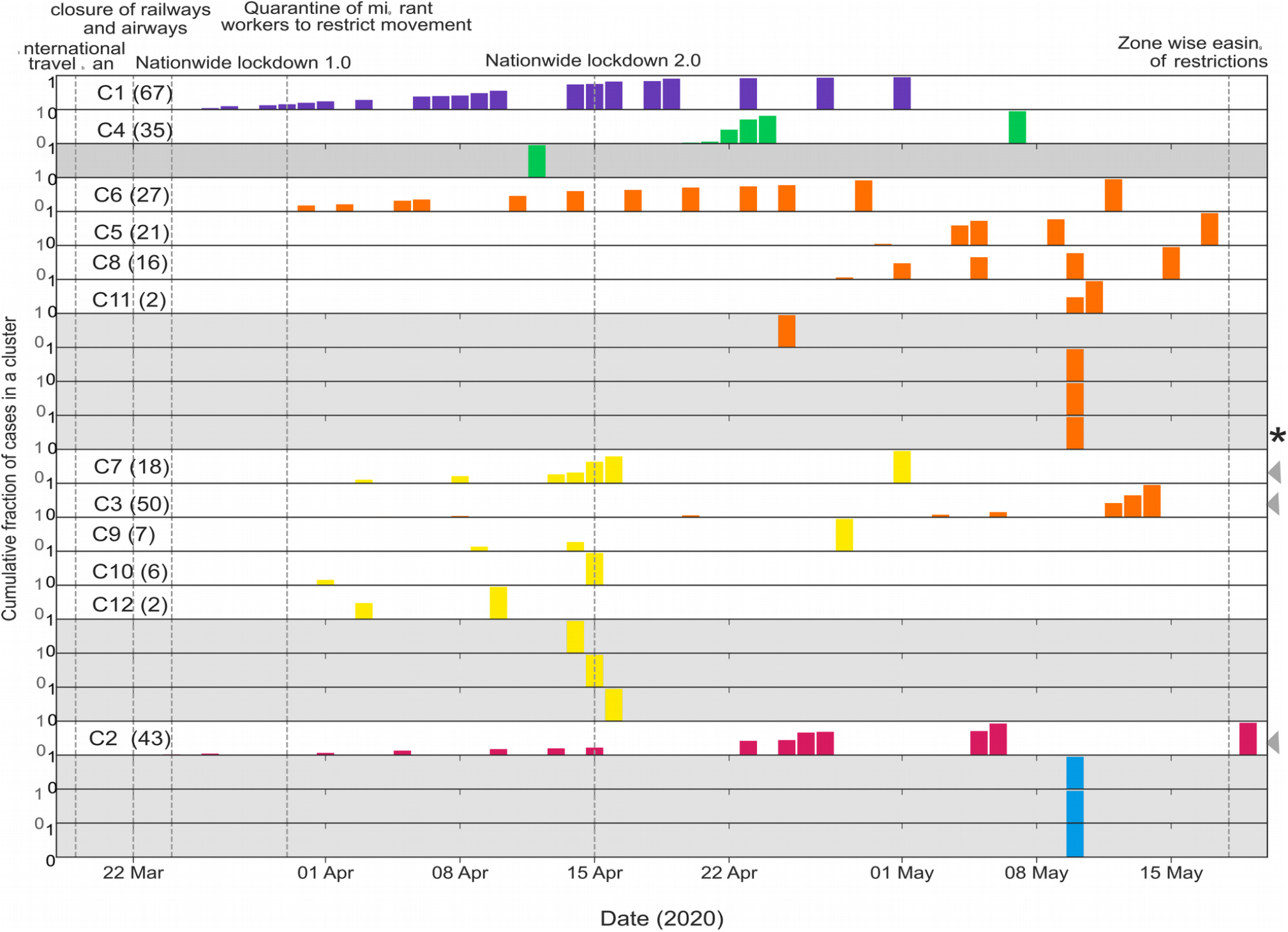
Time course of positive cases in Karnataka by cluster and lineage.X Axis represents time (the duration of the study), y-axis (each row) represents – cumulative fraction of cases (0-1) in a cluster (white) or singleton (grey). Date of interventions are demarcated by vertical lines. Clusters are coloured by majority lineage, mixed Clusters (clusters with more than one lineage) are indicated with an arrow heads on the right. Note: Cluster C1 (purple) and C4 (green) had no new cases since May 1, 2020 and May 7, 2020 respectively. Cases of lineage B/B.6 (orange and yellow) have been detected throughout. Cases of lineage A.p7 (blue) and one of B.6 (indicated by *) were travellers who entered the state close to the indicated time.

## Discussion

SARS-CoV-2, the virus causing COVID19, has now spread throughout the world. Despite restricting travel from affected countries early in the pandemic, India started reporting cases of COVID19 by January 30, 2020 and sustained local transmission was observed in multiple states including Delhi, Maharashtra, and Gujarat^12^. SARS-CoV-2 was first detected in the south Indian state of Karnataka on March 8, 2020 and by May 21, 2020 it had spread to 28 out of the 30 districts of the state resulting in 1578 cases. The data from this study using a combination of genomic epidemiology and contact tracing provides evidence for multiple introductions of the virus into the state, with sustained local transmission. We report the circulation of six lineages of SARS-CoV-2 in the state namely—A.p7, B, B.6, B.1, B.1.1, and B.4 (Pangolin lineage nomenclature). Amongst the 47 virus isolates sequenced in this study, 65.9% (31/47) belong to lineage B and B.6. Most of the contact clusters (9/12) had a single lineage suggestive of multiple introductions of the virus into the state.

Lineage A and B (related to S and L clades of GISAID) of the virus were sequenced in China in January 2020^21^ and they differ at position 8782 in ORF1ab and 28144 in ORF8 respectively. These form the reference sequences and are probably ancestral sequences to other circulating lineages. Viruses from both lineages are now circulating in different countries of the world^21^.

In this study, 3 of the 47 sequences, belong to lineage A.p7 and were from individuals with travel history to other states within India. This lineage is defined by two SNPs T8782C and C28144T and has been reported from Saudi Arabia, Russia, Turkey, and India^21^. No onward transmission was reported from these three cases, however they indicate continued importation of SARS-CoV-2 into the state emphasizing the need for active surveillance of domestic travel.

Of the lineages in the state, B.1 (related to GISAID clade G, and Nextstrain clade A2a) and B.1.1 (related to GISAID clade GR and Nextstrain clade A2a) are European clades. Both lineages harbour the D614G mutation on the Spike protein. It has been suggested that viruses with this mutation are more infectious and the mutation was present at higher frequency in samples across the world^16,24,25^. Of these two lineages, B.1 was a major contributor to the Italian outbreak^21^. In our study, all seven sequenced samples from a large cluster, C4 (comprising of 35 individuals) belonged to this lineage (Figure 3, Supplementary Table 4). This cluster was restricted to Bengaluru city and no new cases were reported from it between 7–21 May. The index case for this cluster was a patient with SARI. Taken together, these observations suggest a hitherto undiscovered link for this cluster to Europe. One other sequence from an individual with no known contact with a positive case in Mysuru district was also assigned to lineage B.1. However, this sequence clustered separately from all the others in the phylogeny and therefore may represent a separate introduction.

The largest cluster in the state, C1 (comprising of 67 individuals) had all three sequenced samples belonging to lineage B.1.1 (Nextstrain clade A2a). Lineage B.1.1 is defined by three additional (to B.1) SNPs—G28881A, G28882A, G28883C^21^. The cluster C1 was initially restricted to Mysuru and subsequently spread to two other districts. Of the 67 cases in this cluster, 16 were symptomatic with the index case being a SARI patient. No new cases could be linked to this cluster after May 1, 2020 up until the conclusion of this analysis (May 21, 2020), suggesting that it had been contained. The index case of this cluster had no history of international travel but was an employee of a company which had a number of international visitors until mid February 2020, including visitors from Europe.

Yet another large cluster (C3) consisting of 60 patients and largely restricted to a quarantine centre in the city of Bengaluru had two lineages B.4 and B. Two of the 47 sequences from C3 were assigned to lineage B.4 (Nextstrain A3). B.4 is a clade associated with travellers from Iran^21^. One sequence from this cluster was from lineage B. This cluster merits further analysis as lineage B.4 was unique to this cluster. The presence of two lineages in this cluster have two possible explanations. Either there were dual introductions of the virus (different lineages) into the cluster or that the two lineages are actually part of two clusters.

Lineages B/B.6 were assigned to the largest number of sequences (31/47) sequences in this study. Using a maximum likelihood based approach we were unable to completely separate B and B.6 as some branches had sequences from both clades (Figure 2). Further, nine of twelve clusters in this study and seven of the eleven cases with no known contact (63.6%) belonged to lineages B/B.6 or both. Lineage B is one of the two clades that were circulating in China in late 2019. Lineage B.6 has earlier been reported from Philippines, UK, North America, Australia, Singapore and has also been reported from other parts of India^15^ (Pangolin). The defining mutations for these lineages are similar to that of the A3i clade which has been described as a distinct phylogenetic group in India^15^. Indeed, upto a third of the cases in multiple states across the country belong to the A3i clade^15^.

These lineages were detected throughout the period in this study and across the state including one domestic traveller who entered the state toward the end of the study period (Figure 4). In Bengaluru city, two clusters C7 and C10 (Supplementary 4), as well as three of eleven epidemiologically unlinked cases were assigned this lineage. Overall, our analysis suggests that the B/B.6 lineage is now established and sustained by local transmission in the state with continued importation from other parts of India.

One of the notable features in this study was the ability to assign virus lineages to cases with no known history of contact with a positive individual. This underscores the utility of genomic epidemiology in filling the gaps of identifying the source of infection.

Some studies had initially proposed a link between viral lineages, transmission and disease phenotypes which have not been substantiated by experimental evidence^26^. The analysis of sequences obtained from symptomatic and asymptomatic (at the time of testing) individuals in this study did not reveal any association with a particular lineage. Symptomatic individuals were spread across lineages B.1, B, and B.6 along with asymptomatic individuals (Figure 2, Supplementary Figure 2). Of the 12 clusters represented in the sequencing data, both the index case and the spreaders were more often symptomatic (Figure 3, Supplementary Table 4). However, sequencing did not reveal any mutations that were specifically associated with clinical state.

Our study had the following limitations – it is a single point analysis and some follow-up data is not available, for instance we do not know if individuals who were asymptomatic at testing later developed symptoms. Further, lineage assignments during an outbreak are dynamic and could change as more data is added and sequencing errors are accounted for. Notwithstanding these limitations, our analysis provides insights about introduction, spread, and establishment of SARS-CoV-2 in Karnataka. Further, we were able to capture both geographic diversity and obtain representation from the ten large contact clusters in the state. This was made possible by linking epidemiological information to genomic data. Integrating such an approach, in real time, into public health measures is essential for an effective outbreak response.

## Data Availability

All complete genomes (47) have been deposited in the GISAID database and are publicly available. The line list of cases is available with the State Surveillance Unit, Directorate of Health and Family Welfare, Karnataka

https://www.epicov.org/

## Acknowledgements

This work would not have been possible without the support of the COVID19 diagnostic team at the Department of Neurovirology, NIMHANS– Ashwini MA, Ruthu Nagraj, Mahesh, Stiben R, Suman Das, Raghavendra Setty TK, Srinivasa R, Tanmoy Nandi, Sourabh Suran, Priti Das, Pallavi SJ, Sathyapriya M, Arpita N Maladkar, Srilatha Marate, Kamala SJ, Gayathri Devi, Kavitha S, Sandhya Rani, Rashmi Kumari, Kumar V, Prasad R, Raja G, Shivakumar V, Jothi Kala C and the data entry team from the State.

We would like to thank the District Surveillance Units and Virus Research and Diagnostic Laboratories (VRDLs) across the state for sample collection, transport and testing. We gratefully acknowledge the contributions of all the laboratories that have submitted their sequences to GISAID, in particular laboratories across India that have been involved in sequencing efforts.

## Author Contributions

PC designed the study, carried out experiments and analysis, interpreted the results and wrote the manuscript, FH designed and carried out the analysis workflow, HPK standardized and carried out experiments, RR carried out experiments/acquired data, VR designed the study, standardized experiments, acquired data (testing), PD acquired, provided access to and curated data, TD curated and entered data, managed line list, NVKR curated and entered data, managed line list, KH curated and entered data, managed line list, AG curated and entered data, managed line list, BJ acquired data (testing and reporting), AP acquired data (testing and reporting), NK curated and filled in missing data, RM acquired data (testing and reporting), revised manuscript, MV acquired data (testing and reporting), SH designed the study, curated data, analysed and interpreted the results, PK Acquired, provided access to and curated data, and interpreted results, AD designed the study, interpreted results and wrote the manuscript, RV designed the study, interpreted results, wrote and approved the manuscript for publication.

## Declaration from the authors

FH is an employee of TrueFactor, In-Mobi group company, however, he was permitted to participate as a volunteer in this study and his employer neither had access to data, nor any say in the design of the study or the decision to publish. All other authors are employees of state (PD, PK) or central government. The employers had no role in the design of the study or the decision to publish. The authors declare that they do not have any other financial or non-financial relationships that could present a conflict of interest.

